# Utility of Technology in the Treatment of Type 1 Diabetes: Current State of the Art and Precision Evidence

**DOI:** 10.1101/2023.04.15.23288624

**Authors:** Laura M. Jacobsen, Jennifer L. Sherr, Elizabeth Considine, Angela Chen, Sarah Peeling, Margo Hulsmans, Sara Charleer, Marzhan Urazbayeva, Mustafa Tosur, Selma Alamarie, Maria J. Redondo, Korey K. Hood, Peter A. Gottlieb, Pieter Gillard, Jessie J. Wong, Irl B. Hirsch, Richard E. Pratley, Lori Laffel, Chantal Mathieu, ADA/EASD Precision Medicine in Diabetes Initiative Consortium

## Abstract

The greatest change in the treatment of people living with type 1 diabetes in the last decade has been the explosion of technology assisting in all aspects of diabetes therapy, from glucose monitoring to insulin delivery and decision making. Through screening of 835 peer-reviewed articles followed by systematic review of 70 of them (focusing on randomized trials and extension studies with ≥50 participants from the past 10 years), we conclude that novel technologies, ranging from continuous glucose monitoring systems, insulin pumps and decision support tools to the most advanced hybrid closed loop systems, improve important measures like HbA1c, time in range, and glycemic variability, while reducing hypoglycemia risk. Several studies included person-reported outcomes, allowing assessment of the burden or benefit of the technology in the lives of those with type 1 diabetes, demonstrating positive results or, at a minimum, no increase in self-care burden compared with standard care. Important limitations of the trials to date are their small size, the scarcity of pre-planned or powered analyses in sub-populations such as children, racial/ethnic minorities, people with advanced complications, and variations in baseline glycemic levels. In addition, confounders including education with device initiation, concomitant behavioral modifications, and frequent contact with the healthcare team are rarely described in enough detail to assess their impact. Our review highlights the potential of technology in the treatment of people living with type 1 diabetes and provides suggestions for optimization of outcomes and areas of further study for precision medicine-directed technology use in type 1 diabetes.

**Preface (Lay Abstract):** We reviewed literature of the last decade to evaluate the impact of technology on the treatment of people living with type 1 diabetes. Screening of 835 articles and in-depth review of 70 showed that novel technologies, ranging from continuous glucose monitoring systems, insulin pumps and decision support tools to the most advanced hybrid closed loop systems, improve important measures like HbA1c and time in range, while reducing hypoglycemia risk. Of importance, several studies showed a positive impact on person-reported outcomes, like quality of life or, at a minimum, no increase in self-care burden compared with standard care.

## Introduction

At the time of the centennial anniversary of the first clinical use of insulin, the treatment of type 1 diabetes has undergone multiple innovations that have advanced the health, well-being, and longevity of people living with the disease.^1^ There have been numerous improvements in insulin formulations, in particular the generation of insulin analogs to create ultra-rapid prandial onset or prolonged basal insulin action aiming at more closely mimicking normal physiology. While these newer insulin preparations helped more people with type 1 diabetes achieve targeted glucose levels, further glycemic improvements required advancements in glucose monitoring technologies. Most recently, factory-calibrated continuous glucose monitors (CGMs) have all but eliminated the need for self-monitoring of blood glucose (SMBG).^2–4^

The discovery and routine availability of hemoglobin A1c (HbA1c) measurements in the 1970s provided for the quantification of overall glycemia, which, in turn, supported the design and implementation of the Diabetes Control and Complications Trial (DCCT).^5^ This landmark study compared intensive insulin therapy, consisting of multiple daily insulin injections (MDI) or continuous subcutaneous insulin infusion (CSII) pump therapy with SMBG, with conventional insulin therapy, consisting of only one to two daily injections of insulin with urine glucose monitoring. The DCCT confirmed the importance of intensive insulin therapy versus conventional insulin therapy on improving glycemic levels and reducing the risk of diabetic nephropathy, retinopathy, and neuropathy as well as long-term adverse cardiovascular outcomes.^5–7^ Since 1993, intensive insulin therapy has remained the mainstay of treatment of type 1 diabetes, albeit with substantial self-care burden placed upon the person living with type 1 diabetes until the creation of advanced technologies that have eased many self-care demands.

This systematic review focuses on these advanced diabetes technologies for the treatment of type 1 diabetes as a part of the second International Consensus Report of the Precision Medicine in Diabetes Initiative (PMDI). The PMDI was established in 2018 by the American Diabetes Association (ADA) in partnership with the European Association for the Study of Diabetes (EASD). The ADA/EASD PMDI includes global thought leaders in precision diabetes medicine who are working to address the burgeoning need for better diabetes prevention and care through precision medicine.^8^

We recognize that type 1 diabetes treatment requires orchestrated education and support around a myriad of activities, including dietary intake, exercise management, insulin administration, glucose monitoring, adjunctive therapies, behavioral health, along with transitions in care across the lifespan, all of which should be tailored to the individual’s needs with a precision medicine approach. Given the rapid evolution of technologies, we have limited our systematic review to the last 10 years of published research on advanced diabetes technologies used in the treatment of type 1 diabetes. While regulatory bodies have recognized glycemic control measured as HbA1c as a proximate outcome of various type 1 diabetes treatments, over the past decade, glycemic outcomes have evolved to include assessment of glucose time in range (TIR) and glucose time below range (TBR) as well as other glucometrics associated with CGM use.^9, 10^ Therefore, we include multiple outcomes in our assessments of technologies for the treatment of type 1 diabetes, including those related to person-reported outcomes (PROs), given the opportunity for technology to mitigate self-care burden. We have sought to evaluate the available evidence to answer the questions as to the 1) efficacy of each technology along with the quality of the evidence, 2) whether individual characteristics can help identify which persons living with type 1 diabetes are likely to derive these therapeutic benefits from the available advanced diabetes technologies, and 3) when in the lifecycle should such approaches be implemented to optimize glycemia, reduce severe hypoglycemia, and preserve health for those living with type 1 diabetes.

## Methods

### Protocol Development

Charged with assessing treatment modalities in people with type 1 diabetes, a group of 13 experts identified key areas of type 1 diabetes management. A protocol for this review was developed and registered on 11/17/2021 at Prospero.com before implementation (https://www.crd.york.ac.uk/prospero/display_record.php?ID=CRD42021271680). Initially, nine areas of interest were identified with topics including adjunctive to insulin therapies, behavioral health, beta cell replacement, exercise, glycemic targets, intensive insulin therapy, nutrition, transition of care, and technology for the management of diabetes (Supplemental Table 1). Recognizing that the biggest change over the past decade has been in technological advances for type 1 diabetes management, our protocol was amended (2/2/2023) to solely focus on this topic.

### Search Strategy, Study Selection, and Screening Process

Using PubMed and EMBASE, Medical Subject Headings (MeSH) and free-text terms (Supplemental Table 2) were used to identify technology-related studies. To assess the sensitivity of the search terms, five key articles were chosen independently, and each was identified through the search.

Abstracts retrieved from the literature search were loaded into the Covidence platform. Eligible studies included individuals with type 1 diabetes or Latent Autoimmune Diabetes in Adults (LADA), randomized controlled trials (RCTs), with a minimum of 50 participants, full text available in English, and published January 1st, 2012 through September 5th, 2022. Secondary papers, extension studies, and PRO-focused studies stemming from RCTs were also included. Full inclusion and exclusion criteria are listed in Supplemental Table 3. Given the rapid evolution of diabetes technologies, the period was restricted to approximate the past decade. The primary goal of the review was to determine whether characteristics of people living with type 1 diabetes across the lifespan could identify the best (tailored) treatment(s) to optimize outcomes, including HbA1c, TIR, weight, hypoglycemia, quality of life, and other measures.

### Data Extraction

At each stage, a minimum of two authors screened titles and abstracts found in the literature search. Disagreements about inclusion were resolved by consensus after consultation with a third reviewer. The next level of screening included reviewing the full text of each publication identified in abstract screening.

Covidence software was used to extract data from eligible full texts by two team members in tandem. A standardized template was made in Covidence to aid with data extraction from each full-text article to document journal citation information, study design and population, limitations, comparison group/controls, intervention, outcomes including subgroup analyses, and limitations.

### Quality Assessment Extraction

Cochrane Risk of Bias (https://methods.cochrane.org/bias/resources/rob-2-revised-cochrane-risk-bias-tool-randomized-trials) was used to assess the quality of the studies as the identified texts were limited to RCTs. Therefore, two team members assessed sequence generation, allocation concealment, blinding of participants/personnel, blinding of outcome assessment, incomplete outcome data, selective reporting, and other sources of bias to determine overall risk of bias. For the purposes of this quality assessment, other sources of bias were deemed present if multiple comparisons were done without appropriate correction, which would lead to high risk of bias, especially in subgroup analyses. If information required to assess quality for each domain was not found in the manuscript, it was coded as “not reported”. GRADE criteria were used to rate the quality of evidence for a given topic.^11^

### Data Analysis and Synthesis

Due to the heterogeneity of technologies used and outcome measures reported, it was not feasible to perform a meta-analysis. Instead, a narrative summary of findings by technology type is presented here.

## Results

A total of 835 studies were screened, of which 70 citations met pre-specified parameters to be included in this report (Figure 1). Of the trials identified, 45 were primary reports of RCT data, 6 reviewed data from an extension phase of an RCT, and 10 were secondary outcome papers. (Figure 2). Studies describing PROs accounted for 9 manuscripts (Figure 2). Quality assessment is visualized in Figure 3 for each of these types of analyses. While the trials were balanced regarding the proportion of females and males, the populations studied were overwhelmingly non-Hispanic white. Of note, over the last decade the technologies studied have shifted towards higher levels of automation (Figure 4).

**Figure 1.**
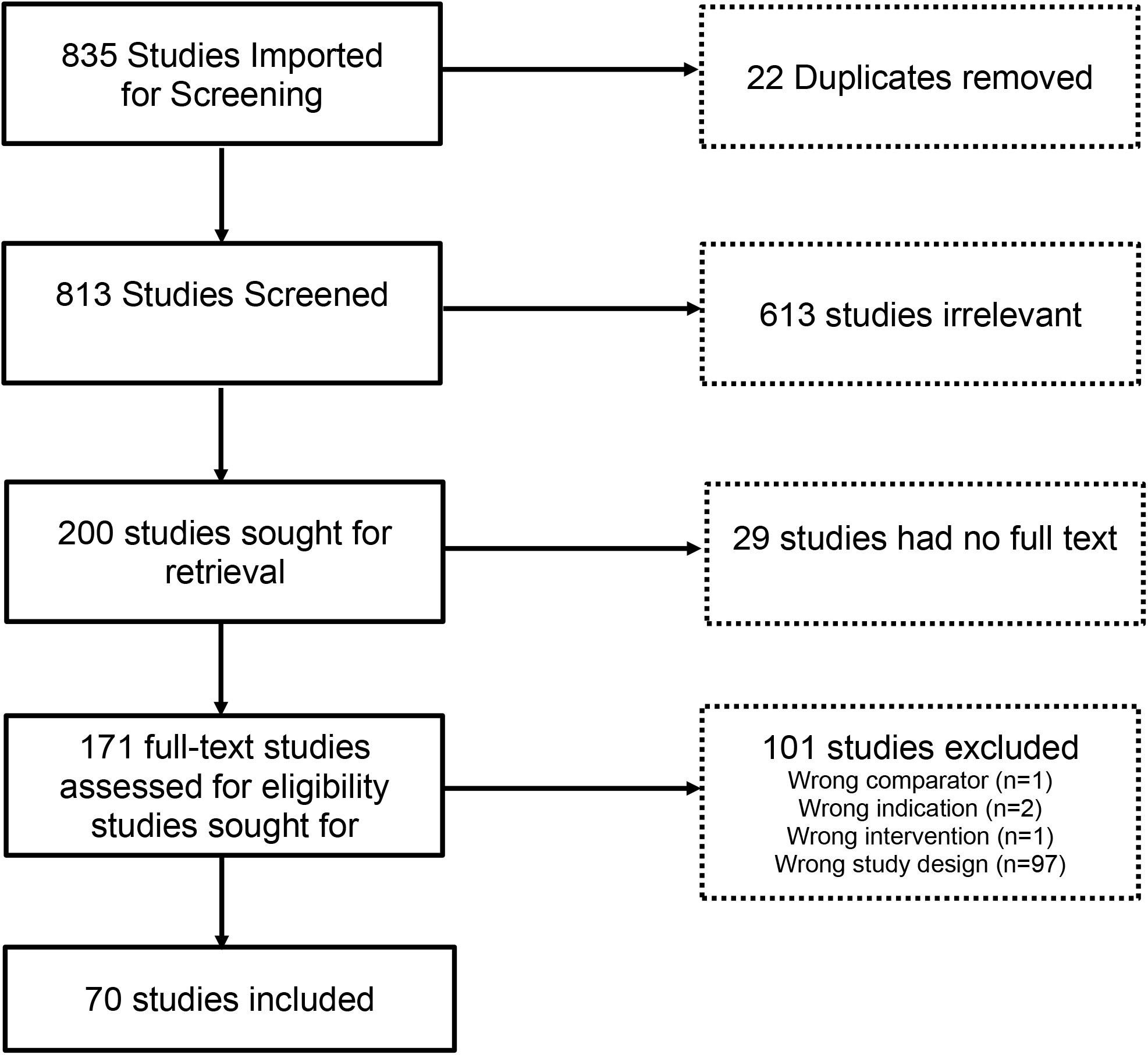
PRISMA flow chart.

**Figure 2.**
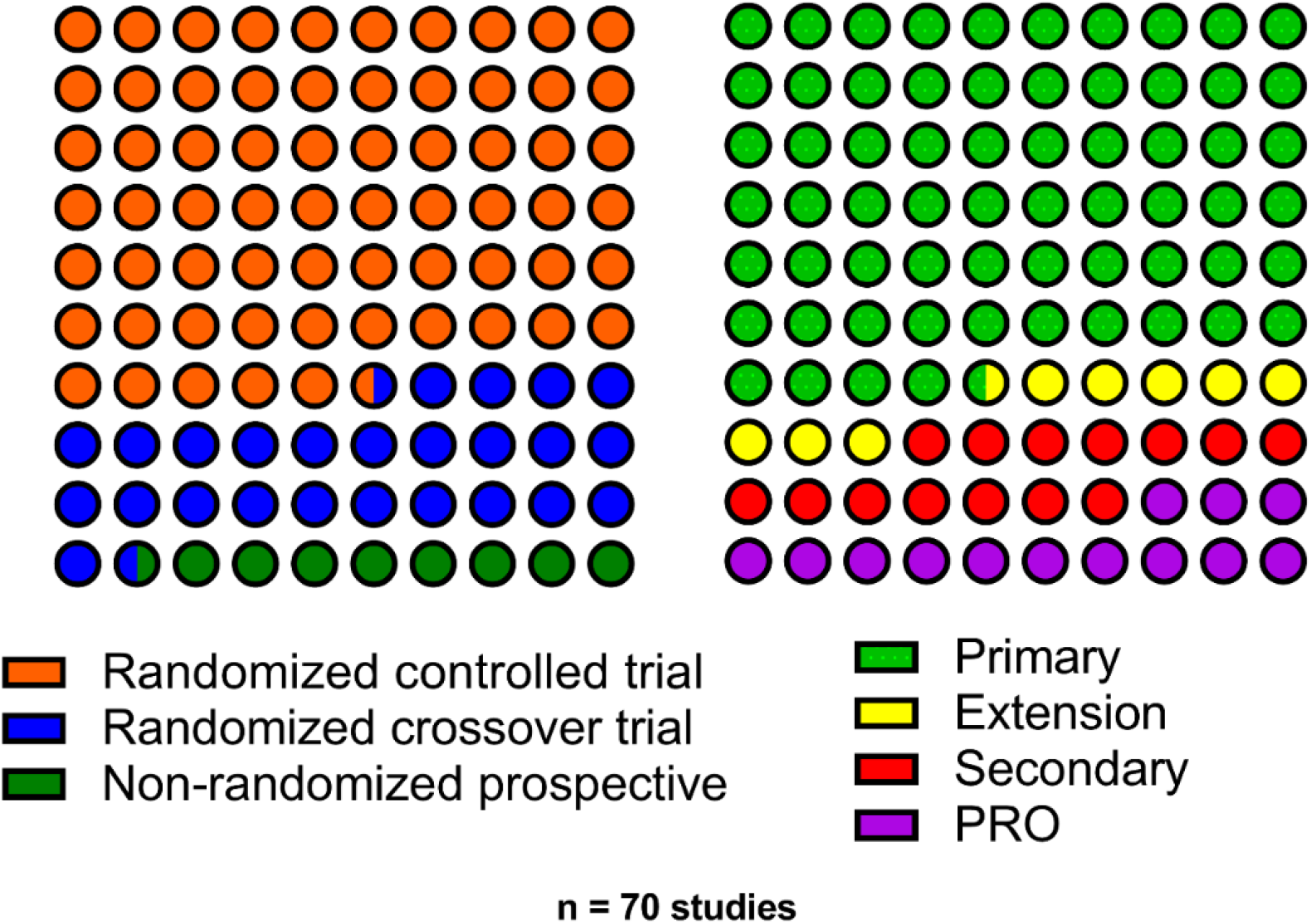
The proportion of studies (n=70) included in the systematic review by clinical trial design (left) and analysis type (right; primary trial results, extension study of an RCT, secondary analysis of an RCT, or person-reported outcome [PRO] studies).

**Figure 3.**
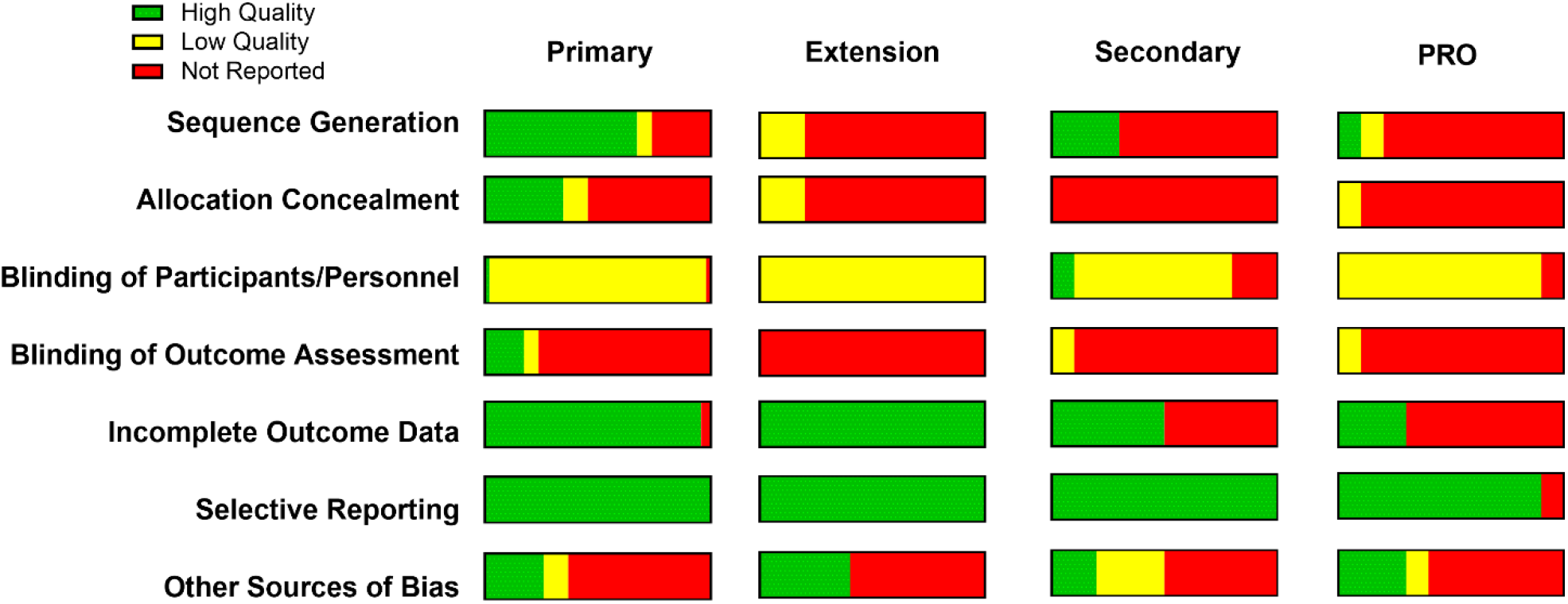
The proportion of studies with a low risk of bias (high quality) or high risk of bias (low quality). This quality assessment is visualized by each individual quality criteria and grouped by the analysis type (primary trial results, extension studies, secondary analyses, or person-reported outcome [PRO] studies). Other sources of bias refer to the presence or absence of a correction for multiple testing.

**Figure 4.**
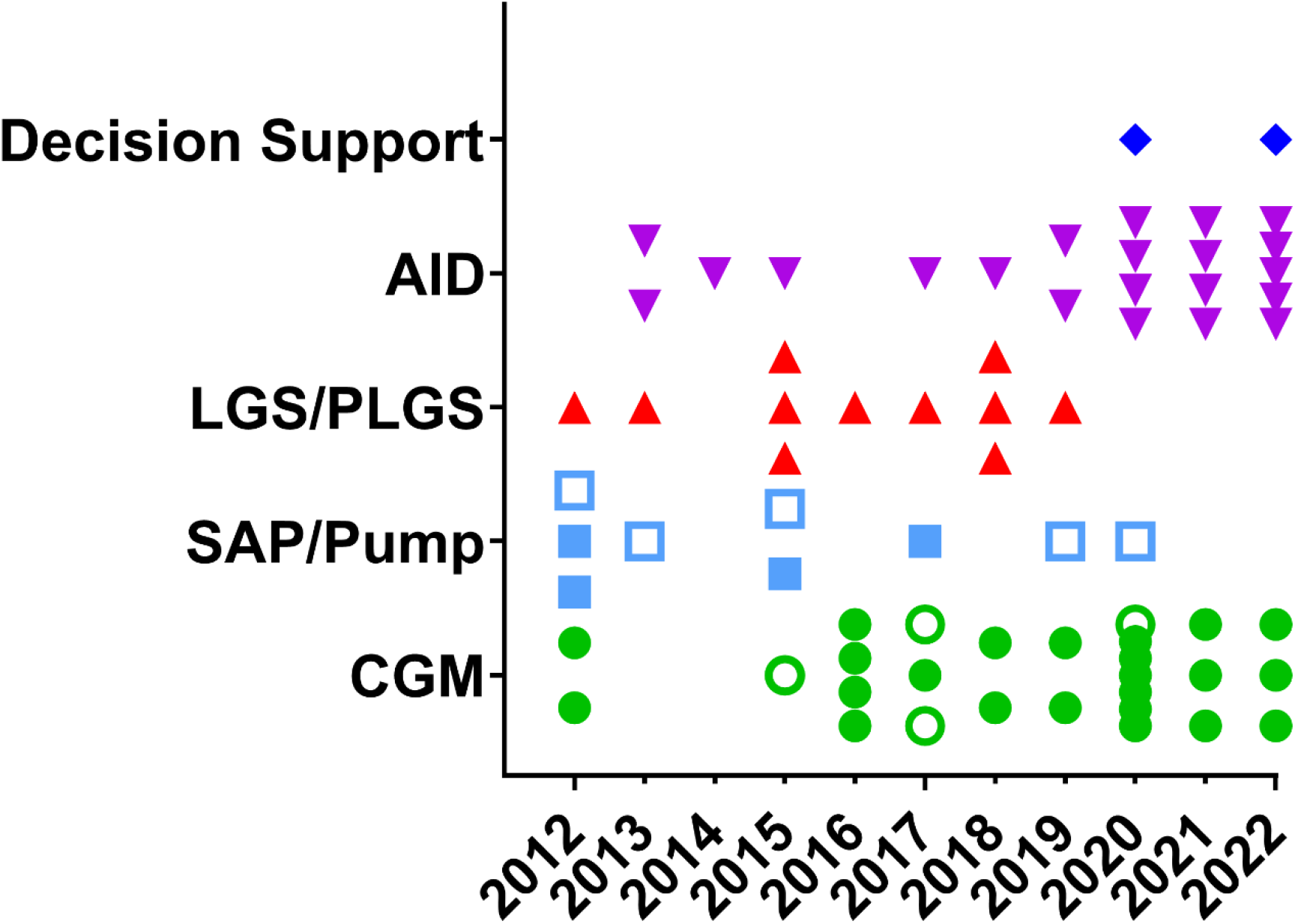
The type of technology studied within each published trial is plotted over time by the year of publication. Each symbol is an individual publication. Open symbols are person-reported outcome (PRO) studies.

### CGM

Technologies allowing continuous measurement of ambient glucose levels in subcutaneous tissue have gained rapid traction in people living with type 1 diabetes. These technologies not only allow insight into glycemic levels without invasive SMBG but provide continuous data with trend analysis and (depending on the systems) alerts of impending hyper- or hypoglycemia. Initially used as diagnostic tools to assist diabetes care teams, they have rapidly become, at least in medium- and high-income countries, the standard of care in glucose measurement for people living with type 1 diabetes. Even prior to regulatory approval, these devices have assisted many people with diabetes in their day-to-day decision making on insulin dosing and provide the foundations for advanced diabetes technologies which tie insulin dosing to sensor glucose levels.

In recent years, many different technologies have become available, with rapid evolution in systems (type of sensor, type of reporting, compatibility of system with smart-phones, integration of reporting in EMR, affordability).^4^ At present, two types of CGM systems are used, depending on the level of user interaction needed to receive sensor glucose information. Real-time CGM (rtCGM) provides updated glycemic information every 1-5 minutes through a continuous connection between the transmitter and receiver, offering real-time alerts. Intermittently scanned CGM (isCGM), also known as flash glucose monitoring, provides the same glycemic information as rtCGM but requires the user to deliberately scan the sensor at least every eight hours to obtain complete glucose data.

As shown in Table 1, the literature search identified 27 articles, 15 primary analyses,^12–26^ 4 secondary analyses,^27–30^ 4 extension studies,^31–34^ and 4 PROs^35–38^. Of primary analyses, 13 were RCTs using parallel (n=10) or crossover (n=3) design comparing CGM to SMBG. The majority of studies investigated standalone-use of rtCGM,^13, 15–18, 20, 21, 27, 30, 31, 33, 34^ while some also examined the use of diagnostic (‘professional’) rtCGM,^14^ do-it-yourself rtCGM,^23^ standalone isCGM,^19, 22, 28, 29^ and decision support systems on top of CGM.^12, 22^ The number of included participants ranged from 52 to 448 with a wide age range (from children to older adults). Three studies had a duration of more than 12 months,^27, 32, 37^ with most studies having a maximum of 6 months of follow-up.^12–25, 28–31, 33, 34, 36–38^

**Table 1:**
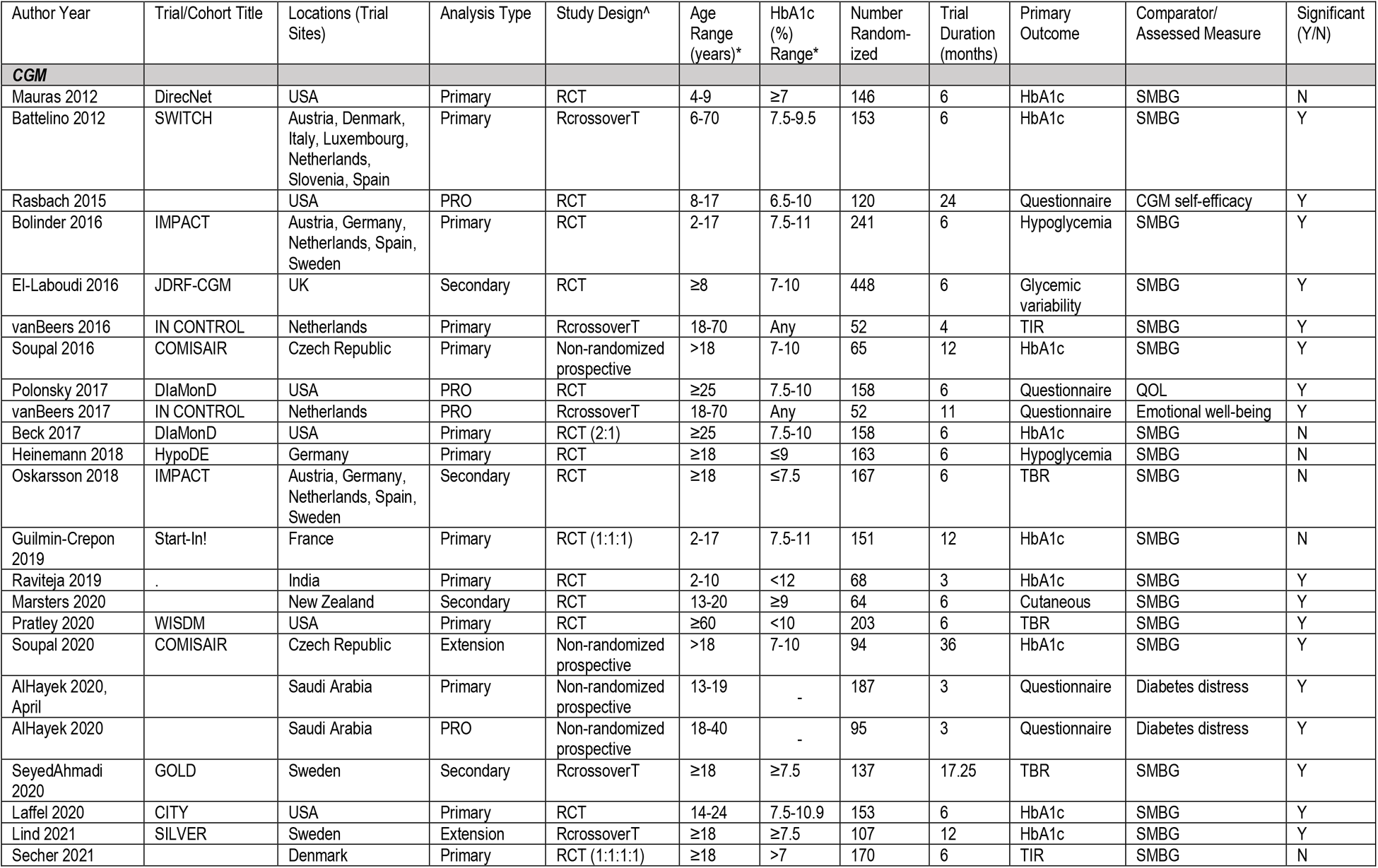
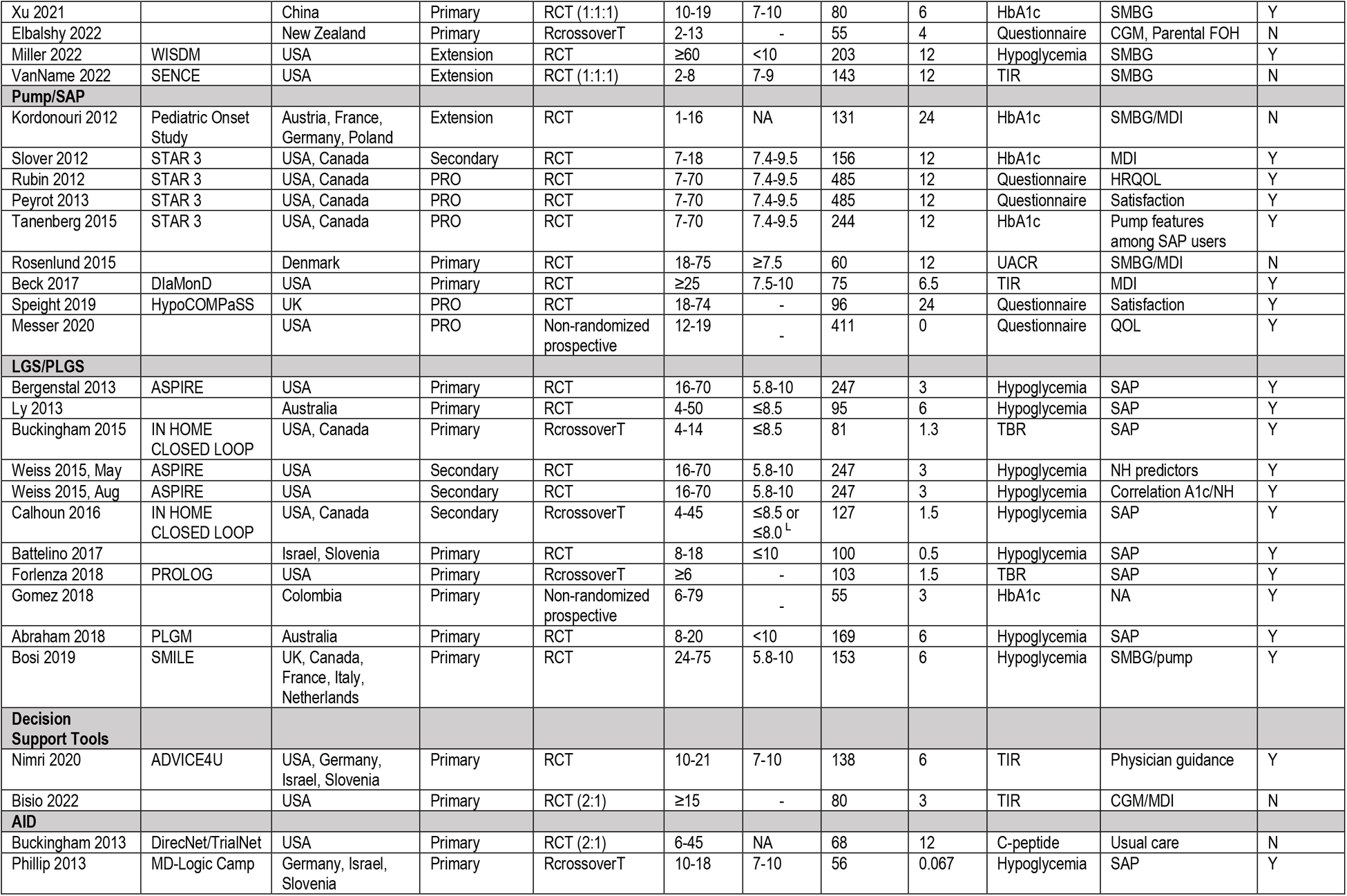
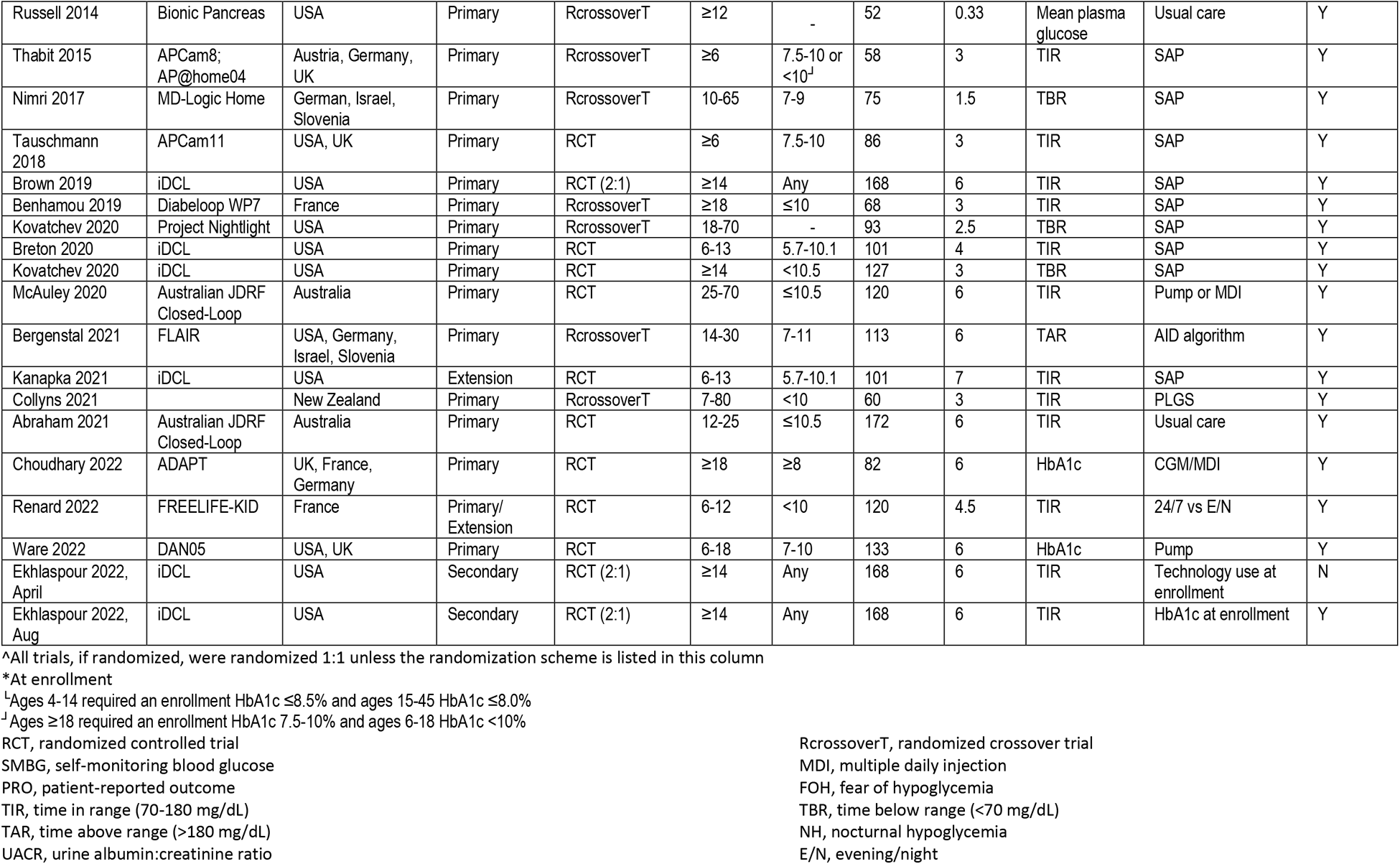
Technology-related clinical trial publications included in the systematic review (2012-2022).

Most RCTs (primary analysis) had HbA1c as the primary outcome,^12, 16, 18, 20, 21, 24, 25, 32, 33^ with hypoglycemia as the primary outcomes in three studies^15, 17, 19^ and TIR^13, 22^ in two studies. One study included the Parental Hypoglycemia Fear Survey as the primary outcome.^23^ Reported secondary outcomes were diverse, encompassing many aspects of diabetes management and well-being (Table 1).

When comparing the method of glucose monitoring (CGM vs. SMBG) half of the RCTs found no statistically significant difference in mean HbA1c,^13, 17–19, 24, 28, 31^ while the other half showed a benefit on HbA1c (mean between-group difference at study end 0.23%-0.60% in favor of CGM).^15, 16, 20, 21, 30, 33^ The most consistent finding was the superior effect of CGM on hypoglycemia prevention and improvement in TIR, which was demonstrated in all but two studies in children.^18, 31^

Of the articles included, 15/27 had low risk of bias for sequence generation and 7/27 had low risk of bias for allocation concealment. Both study personnel and participants were aware of the treatment group in all of the studies. Blinding of outcome assessment could not be assessed in all but 6 papers. The vast majority had complete data and minimal bias for selective reporting. The writing group acknowledges several limitations of the studies evaluating CGM in people living with type 1 diabetes, the short duration of observation, open design of the study, lack of diversity of the populations studied, and the speed of the evolution of technologies (some obsolete by the time of publication). (GRADE evidence: Level B)

Despite rigorous literature search, some articles fulfilling the criteria were not identified. For example, the ALERT1 study prospectively compared isCGM without alerts with rtCGM with alerts in adults living with type 1 diabetes treated with MDI.^39^This six-month study included 254 randomized participants and demonstrated superiority in achievement in TIR, HbA1c, and hypoglycemia risk, including severe hypoglycemia.^39^ Some primary trial results were not identified, like the Strategies to Enhance New CGM Use in Childhood (SENCE) which was a study of 143 children aged 2 to <8 years who were randomized to use of CGM alone, CGM with a familial behavioral intervention, and SMBG that showed no change in TIR but reduced time in hypoglycemia;^40^ yet, an extension of the cohort led by Van Name was included.^31^ A similar issue occurred with the GOLD trial which assessed CGM use in 161 adults with type 1 diabetes on MDI, where the primary study was not included.^41^ Further, publications after our literature search on 9/5/2022 were not included. Focusing on the ALERT1 study, since then the group has published two additional papers. One described an 18-month partial cross-over extension where continued benefits of rtCGM with alarms was demonstrated,^42^ while the other showed that subgroup analysis could not identify any predictive factor differentiating glycemic benefit in people with type 1 diabetes.^43^

### Decision Support

Despite advances in diabetes technologies, for many living with type 1 diabetes, achievement of glycemic targets remains elusive.^44^ Some of this may be due to an insufficient number of providers who can guide diabetes management and the frequency with which follow up is often scheduled. To address this gap and streamline the process of integrating data to optimize insulin doses, decision support systems have been developed in recent years. We identified only two relevant articles reporting primary analysis results of multicenter RCTs on decision support systems. ^45, 46^ These studies randomized 80 and 122 individuals, respectively, and included both adult and pediatric participants. Bisio et al. studied a CGM-based decision support system including real-time dosing advice and retrospective therapy optimization in individuals with type 1 diabetes using MDI over 3 months.^45^ Nimri et al. evaluated an automated artificial intelligence-based decision support system in participants with type 1 diabetes using insulin pump therapy and CGM over 6 months.^46^ The primary outcome was CGM-based TIR for both studies. While Bisio et al failed to show a significant difference in TIR (or HbA1c, analyzed as a secondary outcome),^45^ Nimri et al. demonstrated that TIR in those using the decision support system was not inferior to intensive insulin titration provided by physicians.^46^ Although Nimri et al. also reported statistically significant improvement in HbA1c from baseline to the end of the study in those randomized to the decision support system, there was no between-group difference in HbA1c between the study arms.^46^ Information on sequence generation, allocation concealment, and blinding of outcome assessment were not reported by either of the articles. Both study personnel and participants were aware of the treatment groups. Both articles had complete data and minimal bias for selective reporting. (GRADE Evidence: Level B)

### Insulin Pump and Sensor Augmented Pump Therapy

First tested in the late 1970s, CSII with or without CGM provide a more physiologic means of insulin delivery.^47–49^ Registry data has demonstrated increased use of pumps in clinical practice over time.^44, 50–52^ We identified nine relevant articles meeting our inclusion criteria. Only two articles reported primary analysis results from an RCT.^53, 54^ A secondary analysis of a RCT which focused on the pediatric cohort was also identified.^55^ Another study was an extension analysis that assessed metabolic outcomes one year after the end of the European multicenter randomized Pediatric Onset Study.^56^ The remaining five articles described PROs from RCTs.^57–61^

The samples sizes of these studies ranged from 60 to 485 individuals. The study duration ranged from 6.5 to 24 months, and the primary outcomes were TIR,^53^ HbA1c,^56, 58^ urine albumin-to-creatinine ratio (UACR),^54^ and participant satisfaction.^57, 59, 60^ Beck et al. reported that, in adults with type 1 diabetes using CGM, initiation of insulin pump therapy improved TIR at the expense of increased hypoglycemia and without significant change in HbA1c.^53^ Kordonouri et al. showed that, in participants with sensor-augmented pump (SAP) therapy with frequent CGM sensor use at 24 months, there was significantly less C-peptide loss, though changes in HbA1c were not significant.^56^ Tanenberg et al. demonstrated greater decline in HbA1c with more frequent CGM sensor use in both adult and pediatric participants with type 1 diabetes on SAP.^58^ The next report compared the effect of SAP to MDI on the UACR in individuals with previous or current albuminuria.^54^ In this study, the primary outcome was not met, SAP did not significantly decrease UACR.^54^ Speight et al. reported increased satisfaction regarding “delivery device” and “hypoglycemic control” with insulin pump compared with MDI at 6 months, but this difference did not persist at 2 years.^57^

Information on sequence generation (3/9), allocation concealment (2/9), and blinding of outcome assessment (1/9) was only reported in a few studies. In the nine trials, both study personnel and participants were aware of the treatment groups. Only one study did not have information on selective reporting and 4/9 reported data completeness, all other studies had minimal risk for bias. (GRADE Evidence: Level B)

### Low Glucose Suspend and Predictive Low Glucose Suspend

The ability to pair continuous glucose monitoring with insulin pumps to control insulin infusion rates led initially to two approaches to minimize risk for hypoglycemia.^62^ Low glucose suspend (LGS) algorithms shut off insulin delivery when continuously monitored glucose levels reach a pre-set hypoglycemic threshold. Predictive low glucose suspend (PLGS) algorithms analyze dynamic trends in glucose levels and shut off insulin delivery when hypoglycemia is imminent, but before hypoglycemic thresholds are achieved. Both approaches allow insulin delivery to resume when glucose levels return to pre-specified thresholds. These two approaches, implemented using a variety of CGM and pump platforms, have been tested in many robust RCTs over the last decade.

Among the 11 studies identified,^63–73^ 10 were RCTs, including seven parallel arm comparisons and three with a cross-over design. One non-randomized observational study was identified.^70^ The period of observation varied in the studies from 2 weeks to as long as 12 months. Altogether, 1130 participants were included in the trials, which ranged in size from 55 to 247 participants. Eight reported primary results^63–67, 69–71^ and 3 were secondary analyses.^68, 72, 73^ In 7 of the 8 primary reports, hypoglycemia events or TBR were the primary outcome.^63, 64, 66, 67, 69, 71^ All of these trials demonstrated significant improvements in measures of hypoglycemia with either a PLGS or LGS system.^63, 64, 66, 67, 69, 71^ In one trial, change in HbA1c was the primary outcome, and the PLGS arm was found to be superior to the control group.^70^ Collectively, the findings of these trials indicate that LGS/PLGS results in statistical and clinically significant improvements in risk of hypoglycemia.

Limitations of data include the fact that several different algorithms for suspending insulin were tested, reflecting the rapid technological evolution of the field. As with many other trials in the field, these studies are also limited by the lack of diversity of the populations studied, including diversity in race/ethnicity, access to care, and severity of concomitant diabetes complications and other comorbidities. Of the articles included, 5/11 had low risk of bias for sequence generation and 3 had low risk of bias for allocation concealment. Blinding of outcome assessment could only be assessed in a single study. Most of the trials had complete data and low bias for selective reporting. (GRADE evidence: Level B).

### Automated Insulin Delivery

Building on the strategy of using sensor glucose data to suspend insulin delivery for low or falling glucose levels, developers of automated insulin delivery (AID) systems, also known as closed loop control or artificial pancreas, included programming to increase insulin delivery to mitigate hyperglycemia.^74^ AID systems consist of a glucose sensor, which drives an insulin pump to alter insulin delivery based on algorithmically determined modulations. Early feasibility studies explored use of a fully autonomous system; however, rises in postprandial glucose with this approach led to the use of a “hybrid” approach whereby the user ‘announces’ meals prior to eating. There has been rapid iteration of systems and innovation leading to the testing of multiple pump and sensor combinations with various algorithms forming the backbone of AID.

As demonstrated in Table 1, 21 articles on AID were identified with Figure 4 highlighting the increase in investigations of this technology over time. Eighteen articles reported primary findings from an RCT and encompassed 1,752 participants with individual studies ranging from 52 to 172 participants,^75–92^ two articles providing information on secondary outcomes,^93, 94^ and one from an extension study.^95^ The vast majority of studies had a duration of 3 to 6 months, though some early feasibility studies lasted only a few days^88, 92^ while another study that assessed the impact of using AID shortly after diagnosis followed by SAP on preserving beta-cell function used a year-long intervention.^81^

Based upon the shift that occurred following standardization of metrics to be used for CGM data as well as consensus targets that have been developed,^10, 96^ the primary outcome was TIR in over half the primary trials. Mean adjusted difference in TIR between study groups ranged from 9-15%, which equates to 2 ¼ −3 ½ more hours a day of TIR.^75, 76, 79, 80, 82, 85, 90, 91^

Seminal RCT studies of the Tandem t:slim X2 with Control IQ demonstrated increased TIR in both adolescents and adults^80^ and children aged 6-13 years.^79^ Recognizing that adolescents and young adults often have the greatest difficulty achieving glycemic targets, Bergenstal and colleagues randomized 113 participants aged 14-29 years to a commercially available AID system (Medtronic 670G) followed by an investigational device (advanced hybrid closed loop similar to Medtronic 780G), or vice versa.^77^ Both time above target range (>180mg/dL; TAR) during the day and time <54mg/dL over 24 hours reached statistical significance.^77^ In studies where HbA1c was the primary outcome, whether assessing use of the Medtronic 780G in adults who were previously managed with MDI and isCGM^78^ or in 6-18 year olds using the CamAPS FX AID system,(84) AID systems were found to be superior to the comparator groups.

For quality assessment, sequence generation had a low risk of bias in primary manuscripts (not reported in 9/18). The majority of studies did not provide details on either allocation concealment (11/18) or blinding of outcome assessment (13/18). Participants and study personnel were aware of the randomized intervention as it is not feasible to blind to use of the AID system. Complete data were noted in the trials with minimal bias for selective reporting. (GRADE Evidence: Level B)

Despite the rigorous procedures used, some AID trials that met inclusion criteria were not identified. For example, Ware et al. described use of the CamAPS FX application in children aged 1-7 years old.^97^ In 74 participants, mean age 5.6±1.6 years, undergoing 16-weeks of AID then SAP or vice-versa, TIR rose 8.7% with AID use without increasing TBR, demonstrating the beneficial impact that this AID system can have on very young children.^97^ Additionally, since the last literature search was conducted on 9/5/2022, additional trials have been completed. The pivotal trial describing use of the insulin-only bionic pancreas,^98^ as well as secondary analyses of this cohort, which examined system use in pediatric and adult cohorts,^99, 100^ and results from the extension phase of the trial were published.^101^ Reports on open-source AID systems have documented the efficacy of these do-it-yourself (DIY) systems both in the primary trial^102^ and with continued use during an extension phase.^103^ Two studies have investigated whether attaining strict glucose control through the use of an AID system could preserve residual beta-cell function as measured by C-peptide secretion, both failing to show a difference compared with those receiving standard of care.^104, 105^

Continued exploration regarding use of AID systems in subgroups of individuals living with diabetes, like very young children,^106^ echo the findings regarding the beneficial impact of AID use. For systems initially approved with single-arm studies, post-marketing RCTs have demonstrated benefits of system use.^107^

### Person-reported Outcomes

PROs capture the subjective experience of a person and are obtained through vetted and validated methods such as questionnaires and interviews. PROs offer a layer of understanding of the impact of devices and technologies that goes beyond common glycemic outcomes such as HbA1c and TIR. They are important because even devices that offer glycemic or health benefit can be discontinued by the user if the devices are additionally burdensome or distressing to the person. Further, knowledge of a device’s specific impact on the person’s quality of life offers an additional clinical variable to consider when deciding on type of device to recommend or prescribe. In our review of included PROs, we identified nine studies that had validated PROs measurements.^35–37, 57–61^ In all the cases, PROs served as a secondary (or beyond) endpoint with glycemic outcomes (commonly TIR) as the primary endpoint. Of these nine studies, six were RCTs^37, 57–60^ and two were prospective cohort studies (no control group).^26, 61^ Other studies on PROs were excluded because they were not part of testing of a specific device’s clinical efficacy or effectiveness. The devices tested in these nine studies included CGM, LGS, and SAP. These studies cut across pediatric and adult samples with most focused on those with established diabetes (i.e., few newly diagnosed individuals included).

PROs measured in the studies varied and included self-efficacy about device use, satisfaction with and benefits from the device, diabetes distress, and quality of life. They utilized validated surveys with diabetes distress, one of the most common areas examined. This is consistent with PROs evaluated in studies of people with diabetes—diabetes distress is a common and uncomfortable experience for people with diabetes, and an area that negatively impacts self-management and outcomes. In 5 of the studies, at least one PRO was statistically different from the comparison (control group or non-users of the device) in a favorable direction. Quality of the studies was relatively high given randomized and controlled nature in most cases and validated, commonly used PROs measurements. Participants were aware of the PROs measurements, and they were not asked about specific brands of devices as they completed questionnaires. A number of the studies were funded by the device manufacturer, but the nature of an external ethical review and data management coordinating center kept potential bias low. Overall, evidence from these studies considering the high quality indicates that devices can positively impact PROs. (GRADE Evidence: Level B)

It is not presently known which PRO is most likely to show benefit and if it will happen across all devices and across various subgroups of those living with type 1 diabetes, for example, related to age, duration of diabetes, or previous diabetes management approach. Thus, inclusion of PROs in studies on devices will continue to extend our understanding of the amount, type, and timing of impact to help direct a precision medicine approach to care.

### Precision Assessment

Living with and managing type 1 diabetes is an exquisitely individualized experience that is influenced by many internal and external factors such as age, therapy goals, caregiver needs, and many more. Thus, it is not surprising that the use of technology in the management of type 1 diabetes must also be individualized. The purpose of this systematic review was to assess diabetes technologies using a precision medicine lens. In the previous sections, we critically reviewed the efficacy of technologies and the quality of these clinical trials; herein, we will focus on any precision-based analyses or specific populations studied within these trials and the quality of such analyses.

Of the 70 articles included in our review, 28 (40%) included some form of precision-focused analysis (Table 2)—either post-hoc comparisons of baseline features within or between treatment arms or a trial with a narrowly defined enrollment population (e.g., ages 4-9 years). Baseline characteristics evaluated included age at enrollment, diabetes duration, sex, race/ethnicity, insulin modality (MDI versus CSII), metabolic status (HbA1c, C-peptide, hypoglycemia unawareness status), BMI, presence of nephropathy, education level, and household income. Additional post-treatment variations in efficacy were noted such as differences in the degree of technology engagement or daytime versus nighttime glycemic outcomes. Characterization of these results could benefit people with diabetes and their care teams when seeking to maximize benefit and individualize the approach to technology.

**Table 2:**
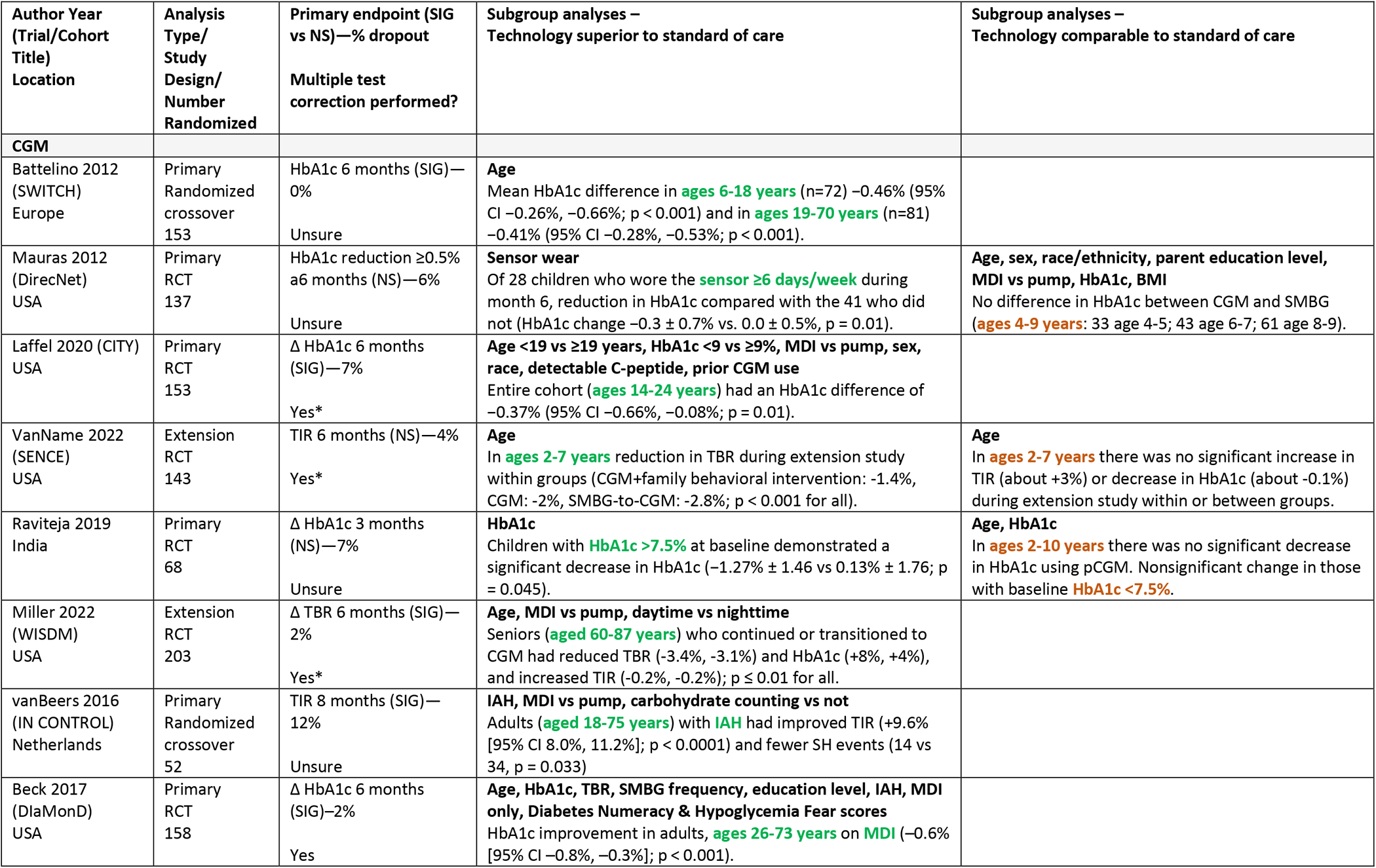
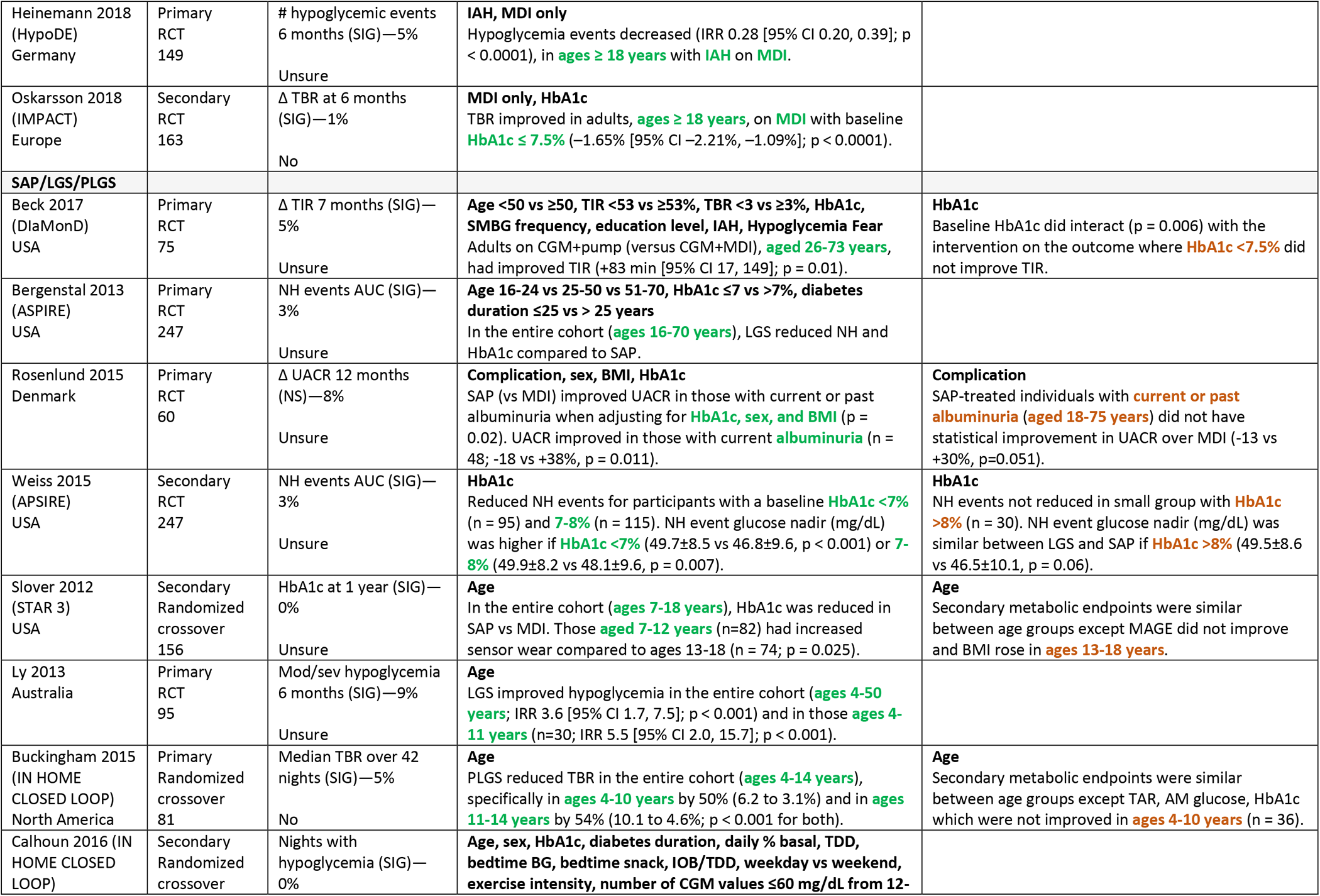
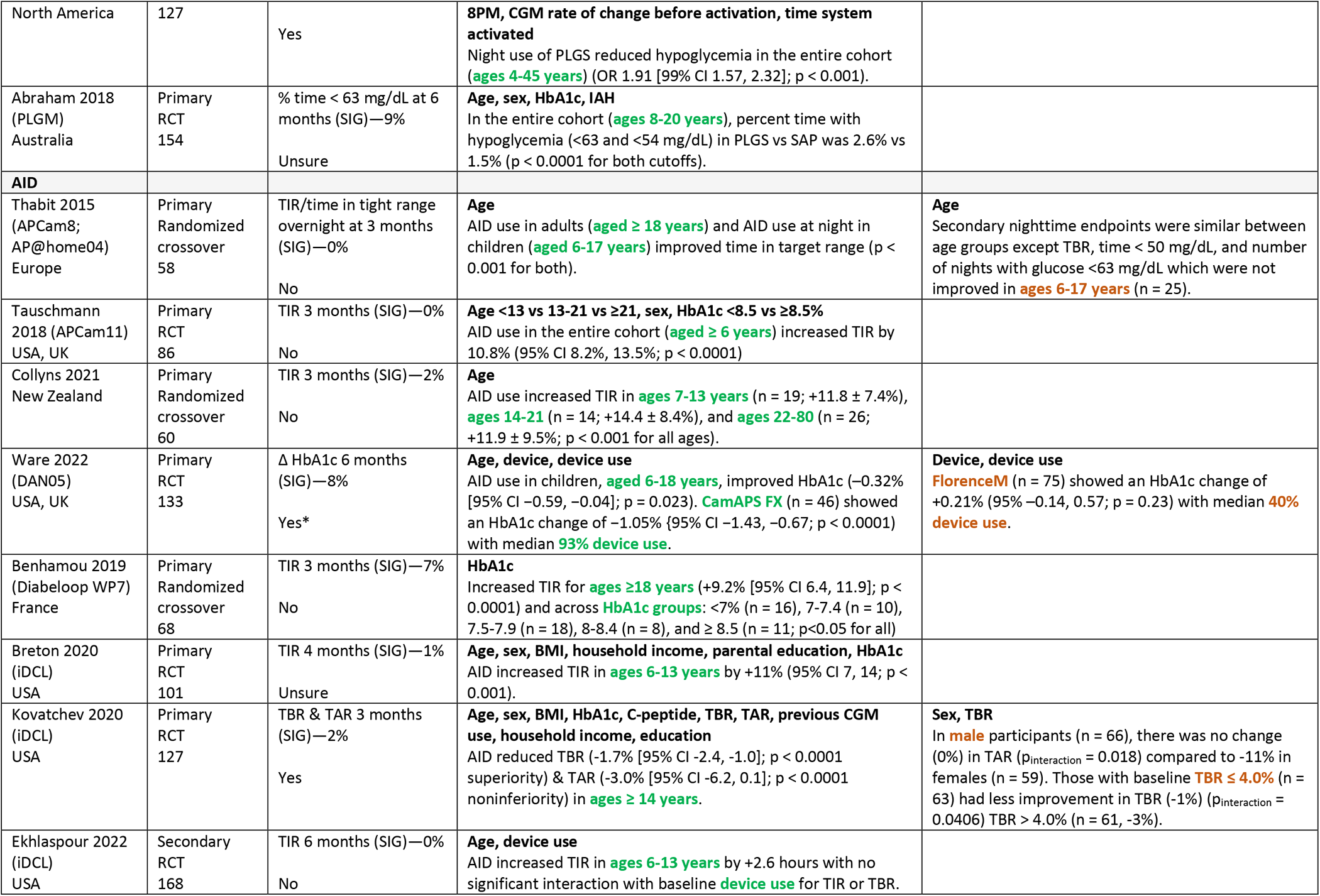
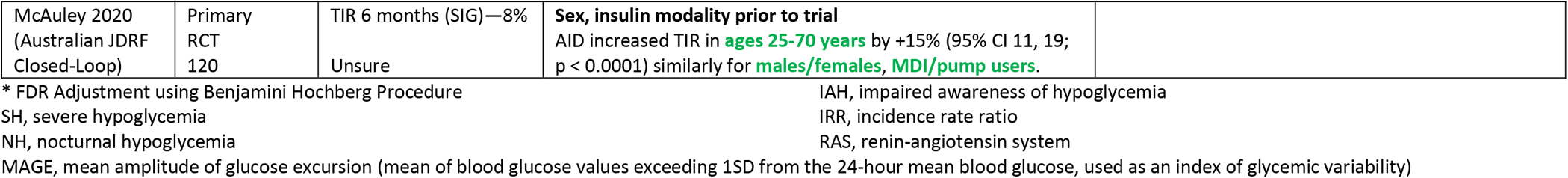
Articles with Precision-Directed Analyses or Subpopulations of Interest

We found the majority of analyses that included pediatric and adult cohorts revealed similar results in metabolic outcomes, though there were some notable exceptions. It is important to remember that these are not direct comparisons of age groups but are post-hoc subgroup analyses of the primary and secondary trial endpoints among those of a specific age range. For example, all cohorts of adults, even stratified within ages 18 to 87 years, had significant metabolic benefits from CGM,^13, 16, 21, 34^ SAP,^17, 53^ LGS/PLGS,^63,71^and AID systems.^76, 82– 85, 90, 91^ Older adults with type 1 diabetes (duration 1-71 years) were studied by Miller et al and had improvements in TBR, TIR, and HbA1c even in the extension phase of the RCT.^34^ Three RCTs in adults using MDI only demonstrated improvement in glycemic metrics with CGM use compared to SMBG.^13, 17, 20, 108^

Technology RCTs in children, especially very young children, however, have produced mixed results. CGM trials in young children, encompassing ages 2-10 years, failed to meet their primary endpoint of HbA1c reduction. The authors posit this could be due to parental fear of hypoglycemia despite overall parental satisfaction with the technology.^14, 24, 31^ Subgroup analyses showed increased sensor wear^24^ or higher baseline HbA1c (>7.5%)^14^ in these trials were associated with improvements in HbA1c with CGM over SMBG. Other endpoints, such as TBR^63, 67^ or hypoglycemia occurrences,^68, 71^ in SAP, LGS, and PLGS trials demonstrated efficacy in children (age range 4-18 years). Adolescents demonstrated worsening of some secondary metabolic endpoints such as increased glycemic variability and increased BMI in an SAP vs MDI trial,^55^ no change in hypoglycemia measures in an AID trial where adults demonstrated improvement,^91^ and males (age 14 years and up) failed to improve TAR while females did.^83^ A consistent picture emerged from trials of AID with efficacy (e.g., improved TIR, TBR, TAR, HbA1c) in all ages (ages 6-70 years) even with age stratification and differences in algorithm and platform.

Other demographic features were sparsely reported, especially race and ethnicity. The diversity of most type 1 diabetes trials, including technology trials, is significantly limited. In pediatric trials, parent education level and income were not significantly associated with outcome but again diversity of these factors may be limited in clinical trials and is an area of need to improve inclusivity.^24, 79, 84^ BMI was only used in stratification or model inclusion in 4 trials^24, 54, 79, 83^ and only affected the outcome in the study by Rosenlund et al in adults with albuminuria.^54^

Baseline HbA1c subgroup analyses in most trials produced consistent results except in 3 studies.^14, 53, 73^ Children with HbA1c >7.5% (vs. <7.5%) on CGM had improved HbA1c at 3 months.^14^ Adults with HbA1c >7.5% (vs. <7.5%) on SAP had improved TIR.^53^ Adolescents and adults with HbA1c <8% (vs. >8%) on LGS had reduced NH events.^73^ The presence or absence of detectable C-peptide did not have any bearing on metabolic outcomes.^16, 84^

Several studies focused on adults with impaired awareness of hypoglycemia (IAH) and demonstrated improvements in TIR, HbA1c, and rates of hypoglycemia with CGM.^13, 17, 20^ In addition, CGM trials often performed subgroup analyses based on the type of insulin modality used by participants, MDI vs pump, and this did not affect the efficacy of any results.^13, 16, 34^ Diabetes Numeracy^20^ and Hypoglycemia Fear^20, 53^ assessment via validated questionnaires did not affect the results of these trials. Unsurprisingly, higher CGM sensor and AID wear time were often tied to increased efficacy.^24, 89, 94^

A special population of adults treated for nephropathy with past or current albuminuria were treated with SAP (vs. MDI) for 1 year and, while there was not a significant improvement in UACR, when these results were adjusted for baseline sex, BMI and HbA1c this became significant.^54^ Additionally, in a subgroup with albuminuria present at screening (UACR ≥30 mg/g), there was an 18% reduction in UACR in the SAP group compared to +38% in the MDI group (*p*=0.011).^54^

These subgroup analyses provide vital, though exploratory, information for people with diabetes and their providers to consider when discussing technology use in type 1 diabetes management. Only 7/28 (25%) included some form of correction for multiple testing and many were not pre-specified analyses. In this age of precision-directed care, more rigorous statistical methods should be considered for subgroups of interest in all RCTs, especially in regard to technology use. In summary, all ages benefit from technology and there are not currently head-to-head comparisons of which device may offer the most benefit for an individual. People with diabetes with IAH or on MDI can still derive significant benefit and should be offered technology. Some technologies may lend themselves better to HbA1c improvement and some hypoglycemia reduction and these can be considered as part of the goals of treatment.

## Discussion

Our multi-disciplinary international team of experts in the management of type 1 diabetes were charged with identifying and analyzing the current evidence regarding a precision medicine approach to the treatment of type 1 diabetes. Given the wide-ranging breadth of treatment approaches to type 1 diabetes, we elected to focus on the modern topic of advanced diabetes technologies and limited our literature review to the past decade. We identified 70 peer-reviewed publications that fulfilled the pre-specified criteria of including people with type 1 diabetes in RCTs with a minimum of 50 participants, published in English on or after January 1st, 2012, and we included any secondary or extension studies from these RCTs. Six non-randomized prospective studies were included from under-represented countries to increase diversity and generalizability. We still uncovered some newer as well as omitted publications germane to our topic, highlighting the dynamic nature of treatment approaches to type 1 diabetes. This collective effort serves to advance the Precision Medicine in Diabetes Initiative, with a major goal of identifying remaining gaps and future directions.

Overall, the studies of advanced diabetes technologies yielded promising results with respect to improvements in glycemia measured as HbA1c or CGM TIR without an increase in hypoglycemia, and often with reductions in hypoglycemia. Many studies also provided positive results with respect to PROs or, at a minimum, no increase in self-care burden compared with standard care. The evidence from this systematic review was considered Grade B in all areas, given the rigorous nature of the studies with notable limitations related to relatively small sample sizes and short durations of follow-up. The assessment of bias was variable across the different areas of diabetes technologies, mostly due to incomplete information regarding sequence generation, allocation concealment, and blinding of outcome assessment although bias was considered low or minimal regarding complete data reporting and selective data reporting.

The DCCT can serve as an example of the evolution of treatment in type 1 diabetes and an approach to precision medicine. Upon its release in 1993, intensive insulin therapy was heralded as the new standard of care for all or most people with type 1 diabetes, although the study was limited to participants aged 13 to 39 years at entry with duration of diabetes of 1 to 15 years. Further, there were other inclusion criteria that would likely limit generalizability to the broader population living with type 1 diabetes. Yet, intensive insulin therapy remained the standard approach, with only more recent recommendations regarding tailoring of glycemic targets to individual needs based upon impaired awareness of hypoglycemia, co-morbidities, or individual characteristics suggestive of reduced survival.^10^

The approach regarding diabetes technology use is reminiscent of the earlier literature on intensive insulin therapy from the DCCT. Following these technology studies, clinical practice recommendations have begun supporting the use of diabetes devices, such as CGM, pump therapy, and even AID systems, in general for people with type 1 diabetes. Many of these technology studies included participants who were more likely to seek advanced diabetes devices, so-called ‘uber’ users. Furthermore, most studies included predominantly white, non-Hispanic participants with higher education and/or socio-economic status from developed countries, with the studies performed by experienced clinical teams often in major diabetes centers that have resources that may not reflect real-world experience.

Similarly, the frequency of contact with clinical teams dictated by study protocols in the DCCT and many technology trials, simply cannot be translated to clinical practice. Reimbursement issues and a limited work force mean people with diabetes often do not have the same support as they attempt more intensive management, as occurred after the DCCT, or integration of technology. Further it is difficult to parse out the impact of education delivered during these touch points and how this influences success with devices.

While these of studies include people with type 1 diabetes from ages 2 to over 70 years of age and most studies included male and female participants nearly equally, there are numerous limitations with regard to other subgroups of potentially salient characteristics such as duration of diabetes, BMI, daily insulin requirements, previous use of diabetes devices, education, co-morbidities, and inclusion of racial/ethnic minorities. Indeed, it has been well-established that disparities exist regarding access to diabetes technologies for those of minority groups; how we overcome these disparities is of critical importance. Thus, there remains a need to evaluate how diabetes technologies should be used within a precision medicine framework regarding subgroups of people with type 1 diabetes, the timing of initiating diabetes devices, requirements for initial training and education, as well as ongoing follow-up and support to sustain technology use. Further, durability of device use also remains an ongoing area for future study.

There is need for the design and evaluation of more real-world experience of the use of advanced diabetes technologies across these many different subgroups and consideration of when the safety of device use needs reconsideration, for example, in an older population where de-prescribing may be warranted or where technology may even have greater benefits. Data generated from registries may provide some insight into device use amongst diverse populations. Registries like the Diabetes Prospective Follow-up initiative, a population-based cohort with individuals from over 500 diabetes centers across Austria, Germany, Luxembourg, and Switzerland, provide data on more than 90% of the population with type 1 diabetes living in those countries. Additionally, leveraging data from these cohorts, it is feasible to explore questions that could never be answered by an RCT. For example, a recent publication noted that use of CGM in youth can reduce the risk of severe hypoglycemia and diabetic ketoacidosis.^109^ Historically, these types of studies would be relegated to GRADE B evidence in the ADA Standards of care, the question becomes whether this data is more relevant to clinical practice as compared to the highly selected populations enrolled in rigorous RCTs.^110, 111^

Whereas there is overwhelming evidence of the benefit of use of technologies in the treatment of people with type 1 diabetes, there also remains substantial need to ensure health equity in the use of advanced diabetes technologies for glycemic and personal benefits across the population of those with type 1 diabetes to reduce the already recognized disparities in diabetes device use. The use of advanced diabetes technologies remains an ongoing challenge in low and middle-income countries. Despite these many challenges, the current state of diabetes technology use in the treatment of type 1 diabetes has revolutionized care with the ability to attain more targeted glycemia, reduced hypoglycemia, and preserved well-being without additional burdens for people living with type 1 diabetes and their families. There is more work to do to understand and then implement a precision medicine model, but progress is underway.

## Data Availability

All data in the present study is from published peer-reviewed manuscripts (systematic review).

## Acknowledgements

We acknowledge several key persons who assisted with the literature search (Krister Aronsson and Maria Björklund from Lund University), instrumental organizational coordination (Chandra Gruber with the American Diabetes Association and Emily Mixter at the University of Chicago), and methodological consultation (Diana Sherifali and Russell deSouza from McMaster University as well as Deirdre Tobias from Harvard School of Medicine).

---

**Supplemental Table 1:** Original type 1 diabetes treatment topics and literature search.

| **Topic** | **Total # of Abstracts screened** |
| --- | --- |
| Adjunctive therapies | 443 |
| Behavioral health | 1665 |
| Beta Cell Replacement | 528 |
| Exercise | 550 |
| Glycemic Targets | 3269 |
| Intensive Insulin therapy | 1181 |
| Nutrition | 699 |
| Transition of Care | 112 |
| Diabetes Technology | 835 |

**Supplemental Table 2:**
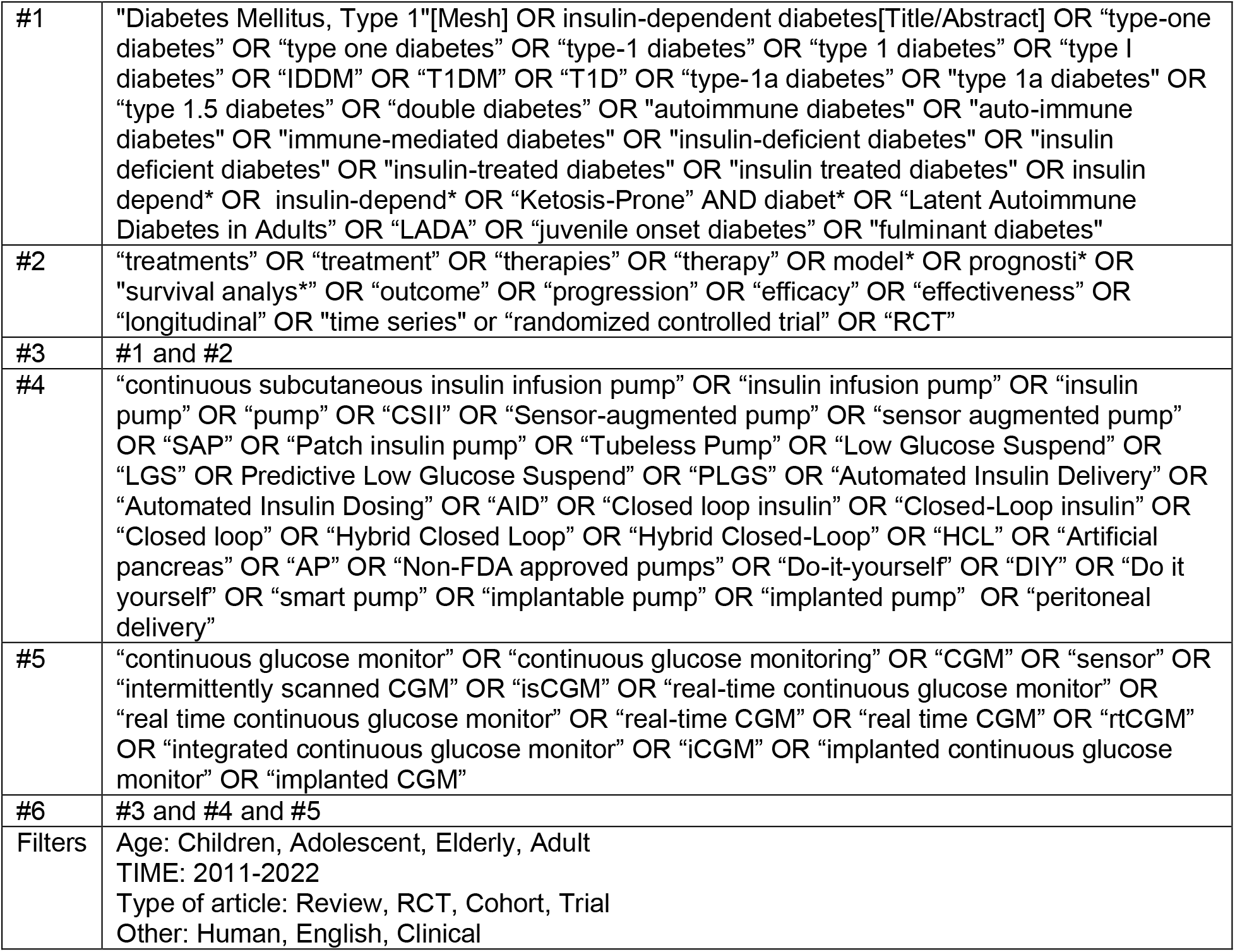
Search strategy – Medical Subject Headings (MeSH) and free-text terms.

**Supplemental Table 3:** Inclusion/Exclusion Criteria for Evidence to Support Recommendations.

| **<u>Included:</u>** | **<u>Excluded:</u>** |
| --- | --- |
| Evidence-level (EL) 1: Randomized controlled trials (RCTs)<br>Human participants<br>English<br>Published, full article in peer-reviewed journal<br>Published January 1, 2012 or later<br>All persons with type 1 diabetes mellitus<br>All persons with LADA<br>Continuous glucose monitoring (CGM), including<br>Real-time CGM<br>Intermittently scanned CGM<br>Integrated CGM<br>Insulin pump therapy, including<br>Continuous subcutaneous insulin infusion<br>Sensor-augmented insulin pump therapy<br>Patch insulin pumps<br>Closed-loop insulin delivery<br>Automated insulin dosing systems<br>Non-FDA-approved/DIY pumps | EL 1 systematic reviews or meta-analyses of RCTs<br>EL 2 studies: meta-analyses including nonrandomized trials or observational studies; controlled trials without randomization; cohort, case-control, cross-sectional studies; epidemiological studies (including surveys and registry data); open-label extension studies; post-hoc analyses.<br>EL 3 network analyses<br>EL 4 consensus/position/policy statements and guidelines<br>EL 3 studies: case reports/series, preclinical/feasibility/protocol/pilot studies, studies with hypothetical cohorts, basic research<br>EL 4 studies: editorials/letters, opinions, reviews, theory<br>Abstracts only<br>N<50<br>Animal studies<br>Non-English<br>Studies published before year 2012, except when cited as background<br>Studies that focus on diabetes technology or treatments that are no longer relevant to practice at the time of publication<br>Studies with a focus on accuracy of a product/device<br>Persons without diabetes<br>Persons with CFRD, type 2 diabetes, gestational diabetes, diabetes in pregnancy, steroid-induced diabetes, monogenic diabetes, MODY, neonatal diabetes<br>Persons with Diabetes s/p chemotherapy, pancreatectomy, post-transplant, medication/drug-induced |

